# Ozone and ultra-fine particle concentrations in a hotel quarantine facility during 222 nm far-UVC air disinfection

**DOI:** 10.1101/2023.09.29.23296366

**Authors:** Petri Kalliomäki, Hamed Sobhani, Phillip Stratton, Kristen K. Coleman, Aditya Srikakulapu, Ross Salawitch, Russell R. Dickerson, Shengwei Zhu, Jelena Srebric, Donald K. Milton

## Abstract

Far-UVC (222 nm UV-C light) is a promising tool to mitigate aerosol transmission of pathogens indoors. However, recent studies have raised concerns related to ozone (O_3_) production and secondary chemistry. In this study, we measured indoor O_3_ and ultra-fine particle (UFP, 17.5-289 nm) concentrations with and without 222 nm far-UVC (average fluence rate 1.7-1.8 µW/cm^2^) in a hotel quarantine facility in Baltimore (MD, USA). We obtained nearby outdoor O_3_ concentrations from the Environmental Protection Agency (EPA) website. In a sealed empty guest room, the average O_3_ concentrations were 3 ppb (UV off, 0.1-0.5 ACH), 16 ppb (UV on, 0.1 ACH) and 9 ppb (UV on, 0.5 ACH). In a standard guest room, the average O_3_ concentrations were 12 ppb (UV off, 1.4 ACH) and 14 ppb (UV on, 1.4 ACH), and correlated with outdoor concentrations (*ρ* = 0.65 – 0.74, p = 2*10^−12^ – 2*10^−29^). A linear regression model, adjusted for outdoor O_3_, estimated that use of far-UVC lamps increased the O_3_ concentration by 5.7 ppb (95% confidence interval (CI) 4.9 – 6.5 ppb) in the standard hotel room. Indoor O_3_ concentrations increased with far-UVC usage, however, the concentrations remained 6-12 ppb lower, on average, than outdoors and well below EPA ambient limits. We did not find a clear relationship between indoor UFP concentrations and UV usage. Although our study was limited by absence of direct outdoor measurements of local O_3_ and UFPs, our findings do not support a major impact of far-UVC on UFP concentrations in the real-world environment that we studied.

## 1. Introduction

Hotels have been used as quarantine facilities to mitigate the spread of severe acute respiratory syndrome coronavirus 2 (SARS-CoV-2) in many countries during the coronavirus disease (COVID-19) pandemic. Although individuals are quarantined in separate rooms, cross infections and intra-hotel clusters have been reported ^1–5^, some leading to community outbreaks and lockdowns^5,6^. Such failed attempts to contain SARS-CoV-2 underscore the importance of adequate ventilation, filtration, and air disinfection to mitigate pandemic respiratory virus transmission.

One effective mitigation measure for aerosol transmission of infections is germicidal ultraviolet (GUV) air disinfection^7–10^. It is a well-established method and has been used for several decades^11^. Traditionally, 254 nm UV light (upper-room GUV)^12-14^ has been the GUV wavelength used in indoor environments. However, usage of 254 nm UV light is typically limited to upper-room or in-duct applications as direct exposure can cause damage to eyes and skin^15-16^ (depending on the UV intensity and duration of the exposure). An alternative GUV wavelength, 222 nm far-UVC, has gained popularity in air disinfection and has been studied intensively, especially since the beginning of the COVID-19 pandemic. It is better tolerated by eyes and skin^17,18^ (compared with conventional 254 nm GUV) and effectively inactivates multiple pathogens, including SARS-CoV-2^19-22^. Due to better tolerance, 222 nm far-UVC can be used in occupied zones of indoor environments and hence has the potential, for example, to reduce close-range transmission more effectively than upper-room GUV.

However, health concerns have been raised related to far-UVC usage. Recent studies have shown that 222 nm far-UVC produces O_3_ and secondary chemistry that can notably affect indoor air quality, particularly in spaces with low ventilation^23–27^. However, these studies are mostly limited to modelling cases and laboratory experiments with only a limited number of studies investigating far-UVC generated O_3_ and ultra-fine particulate matter in real-world settings where far-UVC air disinfection would be recommended as an added layer of protection, such as in hotel quarantine facilities and other high-risk indoor venues^28^. In this preliminary study, we take advantage of ongoing influenza transmission research to examine O_3_ and ultra-fine particle (UFP) concentrations in a hotel quarantine facility with and without 222 nm far-UVC.

## 2. Methods

### 2.1. Research site

This study was performed in a 23-story hotel building in downtown Baltimore (MD, USA). The hotel was used as a quarantine facility during the COVID-19 pandemic, and currently in ongoing influenza transmission research. The hotel has a centralized ventilation system that supplies outdoor air to the hallways and extracts the air through exhaust grilles in the bathrooms. The guest rooms have thermostat and motion sensor-controlled fan coil units for heating or cooling the room air (designed only to recirculate the room air). The fan coil units had MERV 13 filters. All guest room windows were locked and could not be opened to increase ventilation. Two rooms on the 14^th^ floor were used in this study, both equipped with portable HEPA filter units (Breathsmart 45i, Alen, TX, USA, max. CADR 245 CFM). The HEPA units were run constantly on the highest fan speed to ensure good mixing. We used the HEPA filter and fan coil units with and without the filters as part of testing the effect of filtering on particle concentrations.

#### 2.1.1. Sealed unfurnished hotel room

The first room, referred to as the “sealed room”, was a large guest room suite (38 m^2^) on the north side of the building. The room had vinyl flooring and was unfurnished (i.e., no bedding or desks) aside from plastic chairs and tables to place sampling equipment (see Figure 1). The room had a dehumidifier (HomeLabs HME020031N, NY, USA) to control relative humidity (RH, set point at 35%). We sealed door gaps, taped window joints and covered the windows with plastic sheets to ensure minimal ventilation and outdoor infiltration. Hence the room had a low ventilation rate between 0.1-0.5 air changes per hour (ACH), see Supporting Information for more details. The average air temperature and RH during the measurements were 21.1°C and 35.3%, respectively. The room had four 222 nm far-UVC light fixtures (Krypton Shield, Far-UV Inc., MO, USA) each with Krypton-Chloride excimer lamp modules (B1 module, Ushio Inc, Japan). When on, the average fluence rate in the sealed room was modeled to be 1.8 µW/cm^2^ (see Supporting Information for more details).

**Figure 1.**
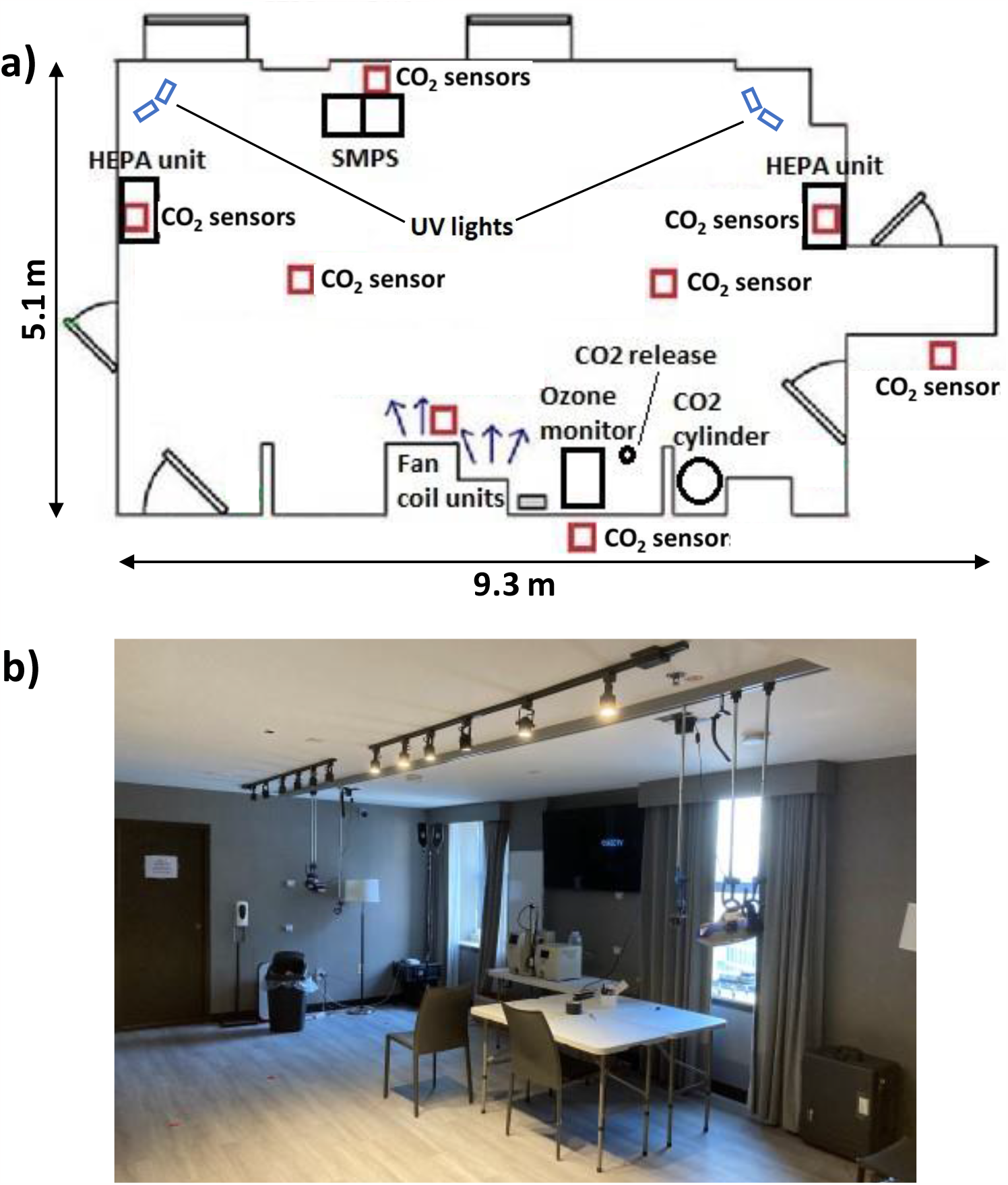
Sealed unfurnished hotel room. a) Floor plan (room height: 2.4 m) and position of far-UVC lamps and sampling equipment. b) Photograph showing SMPS between windows, vinyl flooring, far-UVC lamps and minimal furnishings. Floor plan not in scale.

#### 2.1.2. Standard hotel room

The other room, referred to as the “standard room”, was a furnished single guest room (21 m^2^) on the south side of the building. The room was fully furnished with a king bed, two desks, and carpet flooring (see Figure 2). We used the room under typical operating conditions and did not seal window joints, door gaps, etc. The average air exchange rate of the room was 1.4 ACH and it varied between 1.2 to 1.7 ACH (see Supporting Information for more details). The average air temperature and RH during the measurements were 22.8°C and 23.6%, respectively. The room had three 222 nm far-UVC light fixtures (Krypton Shield, Far-UV Inc., MO, USA), two with Ushio B1 modules and one with B1.5 module (Ushio Inc, Japan). When on, the average UV fluence rate in the standard room was modeled to be 1.7 µW/cm^2^ (see Supporting Information for more details).

**Figure 2.**
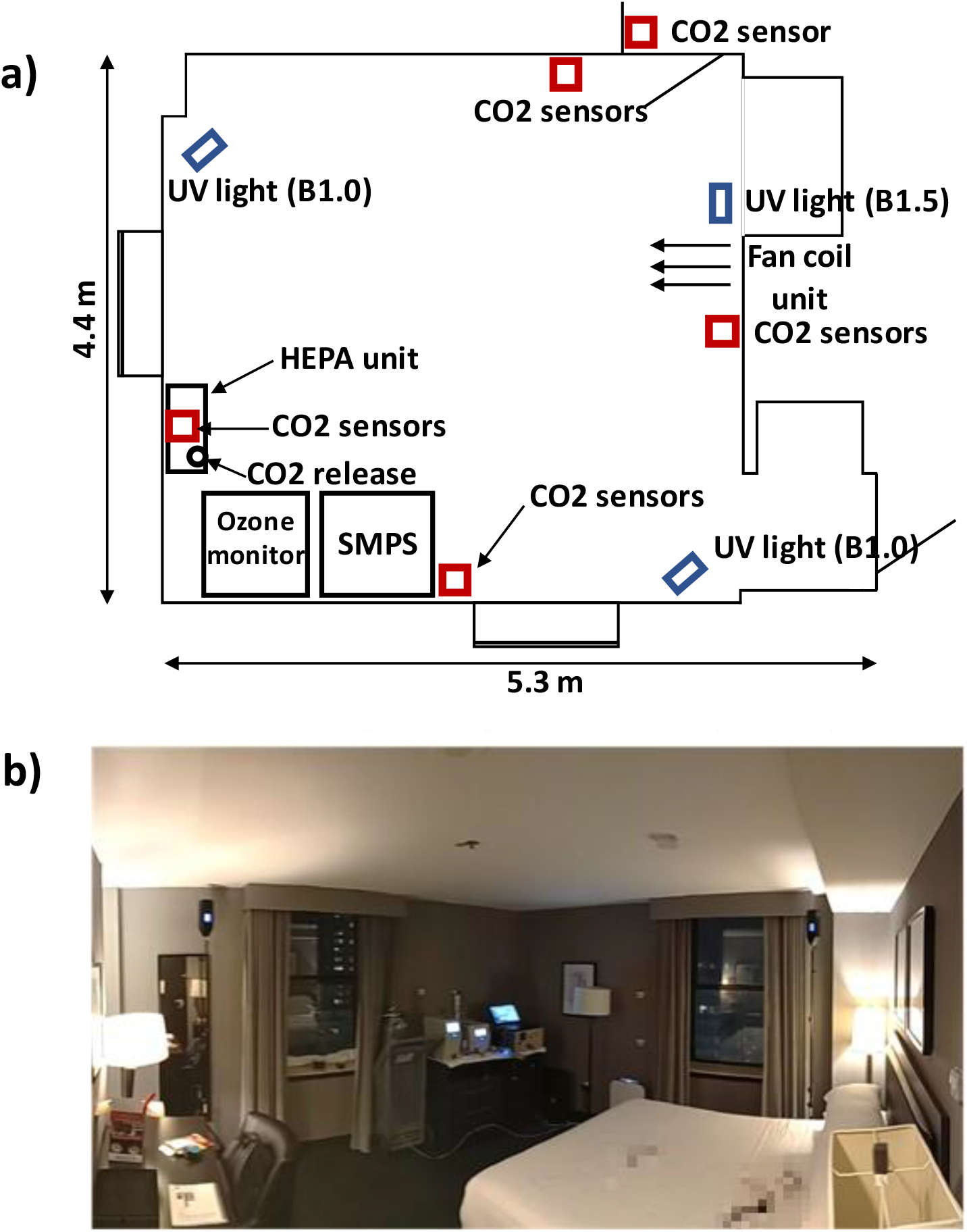
Standard hotel room. a) Floor plan (room height: 2.4 m) and position of far-UVC lamps and sampling equipment. b) Photograph showing far-UVC lamps, sampling equipment, and standard furnishing. Floor plan not in scale.

### 2.2. Measurements

#### 2.2.1. O_3_ measurements

We used an O_3_ monitor (TE49C, Thermo Environmental Instruments, MA, USA) to measure O_3_ concentrations inside the rooms. The concentrations were measured with 10-s resolution and later averaged over 1-min and 1-h intervals. The instrument was calibrated against a reference monitor (Model TE49i-PS Ozone Primary Standard) that was compared to a reference instrument at the National Institute of Standards and Technology (NIST, USA). The absolute accuracy of the instrument is approximately 4 ppb at 10-s resolution and 2 ppb for hourly averages (95% Confidence Interval (CI)).

Outdoor O_3_ concentration data (1-h resolution) was downloaded from the Environmental Protection Agency (EPA) website (https://aqs.epa.gov/aqsweb/documents/data_api.html). We included data from the three closest monitoring stations (12-19 km from the hotel) around the Baltimore metropolitan area in our analysis (which had the highest data coverage during the time of the measurements). Average O_3_ concentration (of the three outdoor locations) was used in the final analyses.

#### 2.2.2. Ultra-fine particle measurements

We used a scanning mobility particle sizer (SMPS 3938, DMA 3081A, CPC 3756, TSI Inc., MN, USA) to monitor UFP concentrations within the size range of 17.5-289 nm. The sampling resolution of the SMPS system was 66 s.

#### 2.2.3. CO_2_ measurements

We monitored ventilation by releasing carbon dioxide (CO_2_) into the rooms at a constant 0.4 L/min rate (GFC171S mass flow controller, Aalborg Instruments & Controls Inc., NY, USA). We used wireless sensors (Aranet4, SAF Tehnika JSC, Latvia) to measure CO_2_ concentrations in the rooms, hallways and outdoors (1-min resolution). The sensors were calibrated with a standard CO_2_ gas with a range of concentrations between 500-10,000 ppm. The CO_2_ sensor accuracy is ±30 ppm or ±3% of the reading (according to the manufacturer). The sensors also measured ambient temperature, RH, and barometric pressure.

### 2.3. Data collection and analysis

We carried out measurements in the sealed room between Feb 17^th^ – Feb 20^th^, 2023 (O_3_ between Feb 16^th^-20^th^, 2023) and in the standard room between Feb 27^th^ – Mar 10^th^, 2023. All data were cleaned (occupied periods removed from the data, etc.) and analyzed with MATLAB. One-hour averages of the data were used in final analyses. Linear regression models were calculated using fitlm-function in MATLAB.

## 3. Results

### 3.1. CO_2_ and O_3_ concentrations in the sealed hotel room

CO_2_ and O_3_ concentrations in the sealed room are shown in Figure 3. Constant injection of 0.4 L/min CO_2_ produced two steady-state levels with concentrations of c. 1060 ppm and c. 2400 ppm. We observed that the fan coil units were pulling air into the room from the pipe chase. Consequently, the average air exchange rate in the sealed room was calculated to be 0.1 ACH and 0.5 ACH when the fan coil units were off and on, respectively. We did not see a similar effect in the standard room.

**Figure 3.**
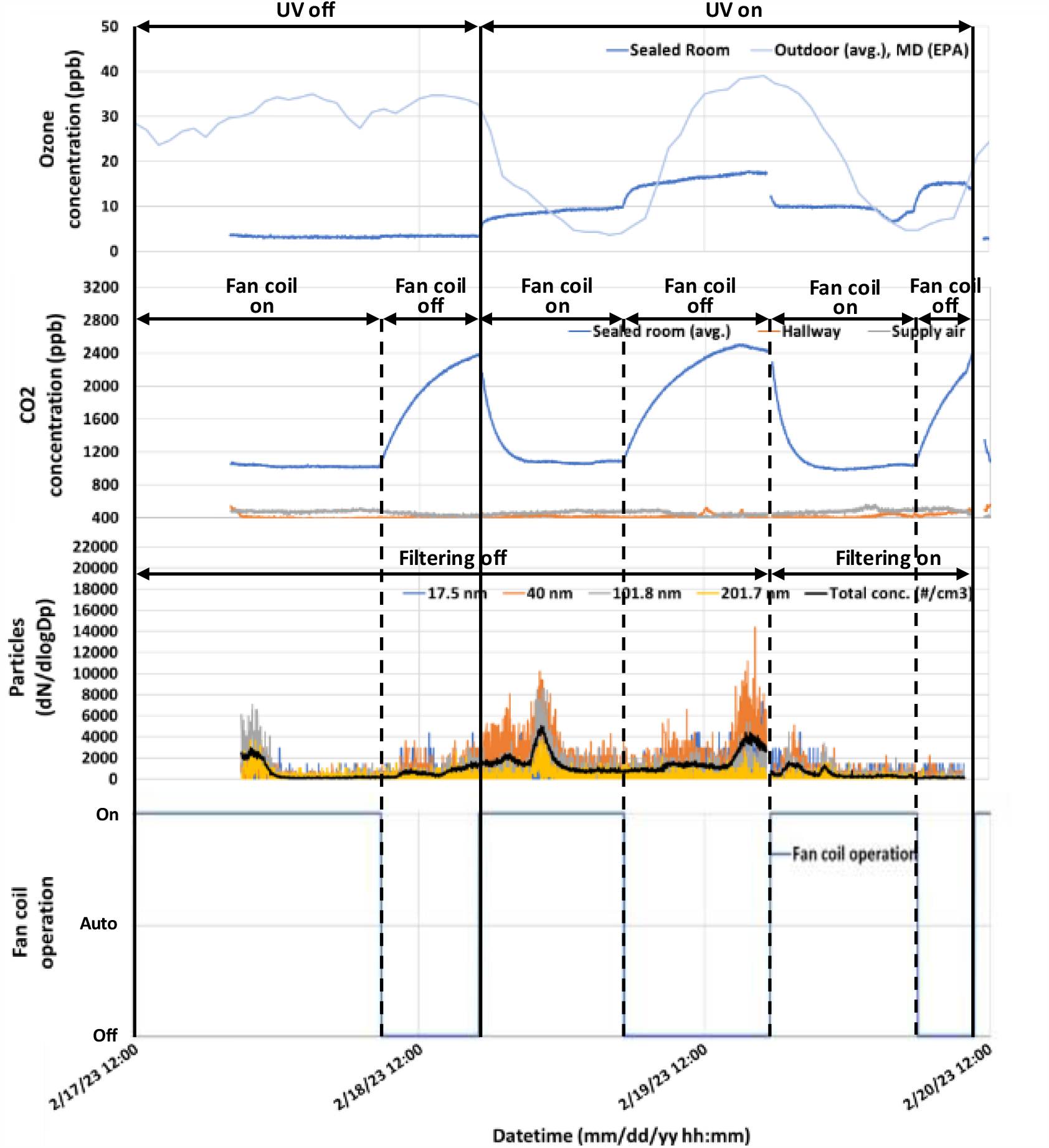
Sealed room O_3_, CO_2_ and ultra-fine particle measurements plotted against time. Periods with far-UVC lamps (on/off), in-room HEPA filtering (on/off) and fan coil unit states (on/off/auto) are shown. Fan coil unit had 2 states: off (0.1 ACH) and on (0.5 ACH). During the filtering periods the fan coil unit had a MERV 13 filter and the HEPA filter unit had a HEPA filter but they had no filter during other times. The outdoor O_3_ levels are an average of three Maryland sites (Essex, Glen Burnie, Padonia) in the EPA Ambient Air Monitoring Network. The CO_2_ concentration in the sealed room is an average of all sensors in the room.

When the far-UVC lamps were off, the average O_3_ concentration was 3 ppb in the sealed room and 32 ppb outdoors at 0.1 ACH (fan coils off), and 3 ppb indoors and 29 ppb outdoors at 0.5 ACH (fan coils on). When the UV lamps were on, the average O_3_ concentration was 16 ppb indoors and 22 ppb outdoors at 0.1 ACH (fan coil units off), and 9 ppb indoors and 16 ppb outdoors at 0.5 ACH (fan coil units on). In general, O_3_ concentrations in the sealed room showed a clear build-up and decay in relationship to operation of the far-UVC lamps and the fan coil units.

Indoor O_3_ concentrations in the sealed room (at 0.1 ACH) were strongly correlated (Spearman’s rank correlation *ρ*) with outdoor concentrations (Figure 4a) both with far-UVC lamps off (*ρ* = 0.88, p = 7*10^−9^) and on (*ρ* = 0.97, p = 3*10^−10^). When the fan coil units were on and the ventilation rate increased to 0.5 ACH, indoor O_3_ was weakly to moderately correlated with outdoor concentrations when the far-UVC lights were off (*ρ* = 0.48, p = 0.04) or on (*ρ* = 0.38, p = 0.06) (Figure 4b).

**Figure 4.**
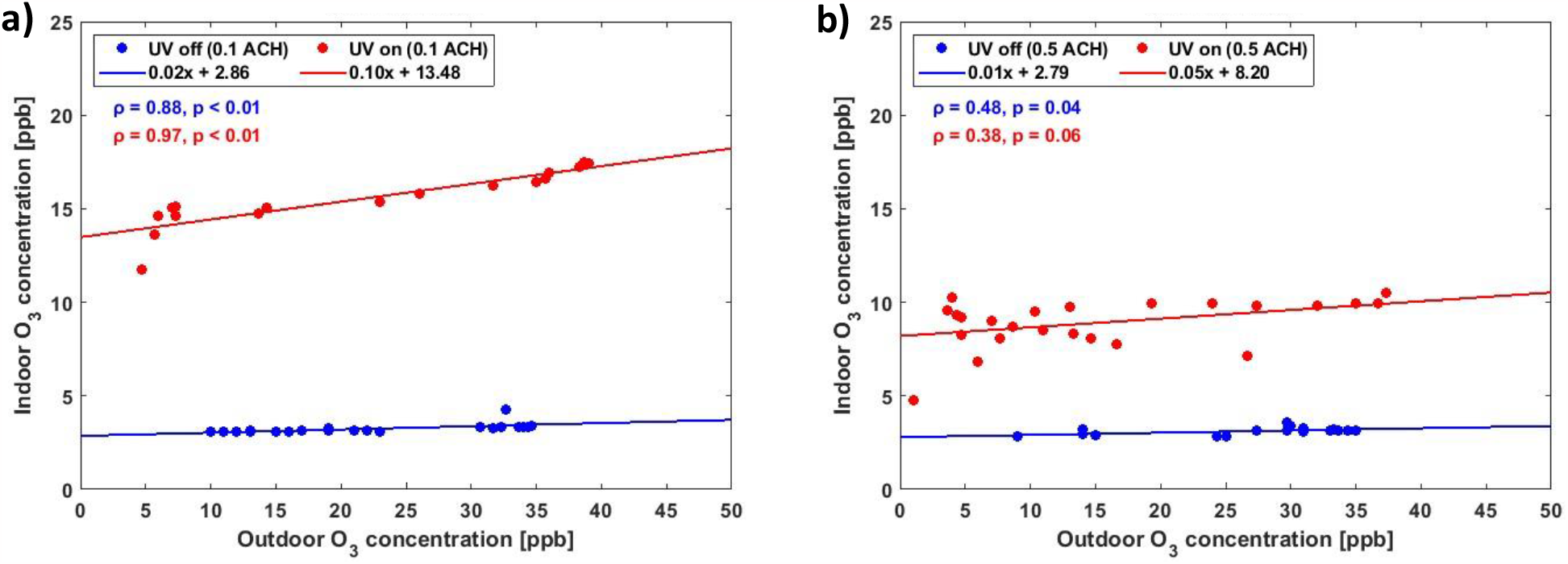
Sealed room indoor-outdoor concentration correlations with and without UV with (a) 0.1 ACH (fan coil off) and (b) 0.5 ACH (fan coil on) ventilation rates. The outdoor O_3_ levels are an average of three Maryland sites (Essex, Glen Burnie, Padonia) in the EPA Ambient Air Monitoring Network. *ρ* is the Spearman’s rank correlation coefficient.

### 3.2. CO_2_ and O_3_ concentrations in the standard hotel room

CO_2_ and O_3_ data collected from the standard hotel room is shown in Figure 5. The average CO_2_ concentration was 840 ppm with notable fluctuations (range: 720 ppm to 1010 ppm). The fluctuations seemed to be driven by changing ventilation due to outdoor infiltration and occasional entry of researchers to check and operate equipment.

**Figure 5.**
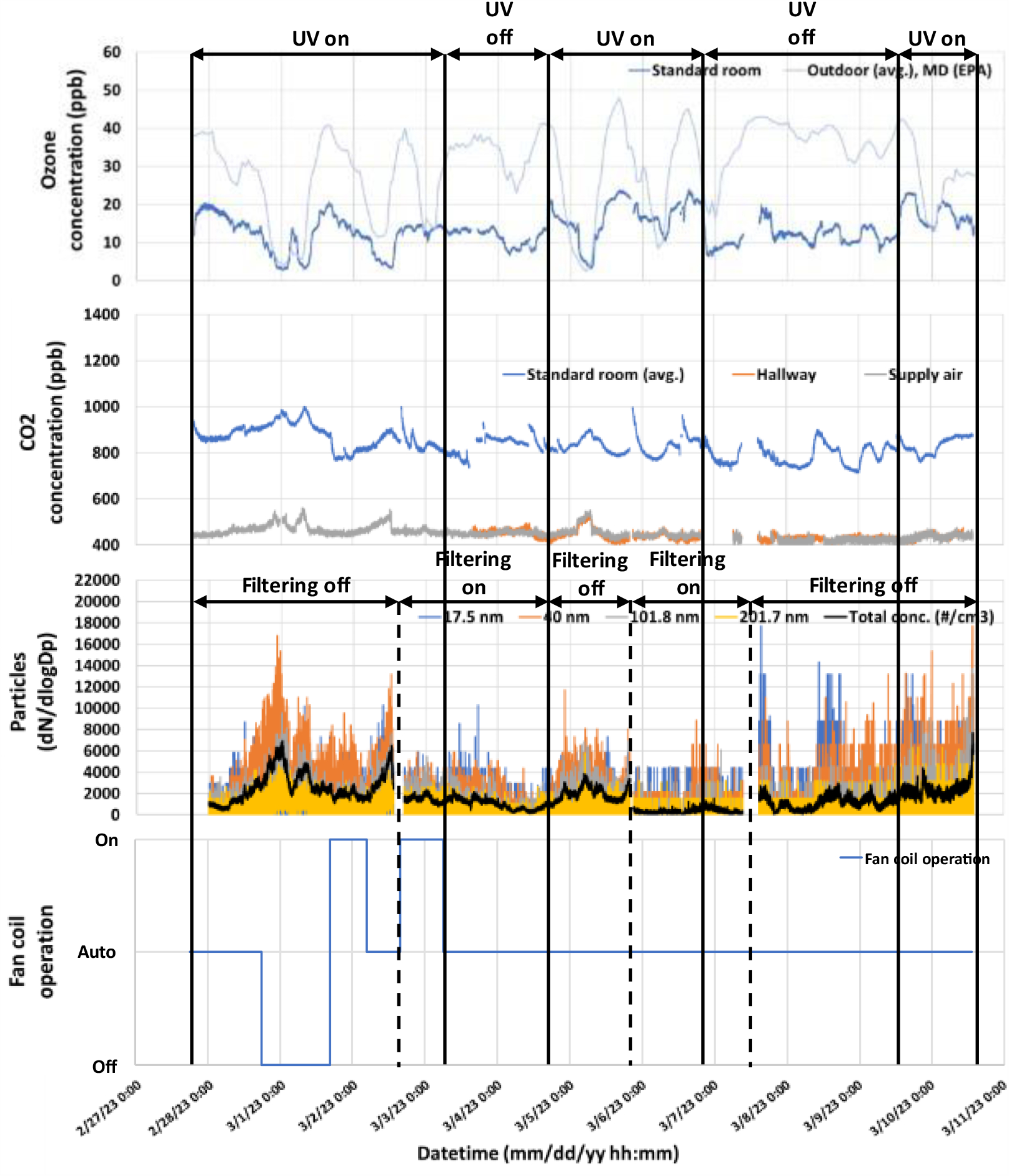
Standard hotel room O_3_, CO_2_, and UFP measurements from February 27 through March 11, 2023. Periods with far-UVC lamps and in-room HEPA filtering on and off and fan coil unit states (on, off and auto) are shown. During the auto state, the fan coil unit ran intermittently as controlled by a thermostat and motion sensor. During the filtering periods the fan coil unit had a MERV 13 filter and the HEPA filter unit had a HEPA filter but they had no filter during other times.

The average indoor O_3_ concentration was 12 ppb when the far-UVC lamps were off and 14 ppb when the UV lamps were on. Average outdoor concentrations were 35 ppb and 26 ppb when the UV lamps were off and on, respectively. Indoor O_3_ was moderately to strongly correlated with outdoor O_3_ concentrations (Figure 6) when the UV lamps were off (*ρ* = 0.65, p = 2*10^−12^) or on (*ρ* = 0.74, p = 2*10^−29^). A linear regression model, adjusted for outdoor O_3_ concentration, indicated that the usage of the UV lamps increased the indoor O_3_ concentration by 5.7 ppb (95% confidence interval (CI) 4.9 – 6.5 ppb). A 1-ppb increase in outdoor O_3_ was associated with a 0.33-ppb increase in indoor O_3_ (95% CI 0.29 – 0.36).

**Figure 6.**
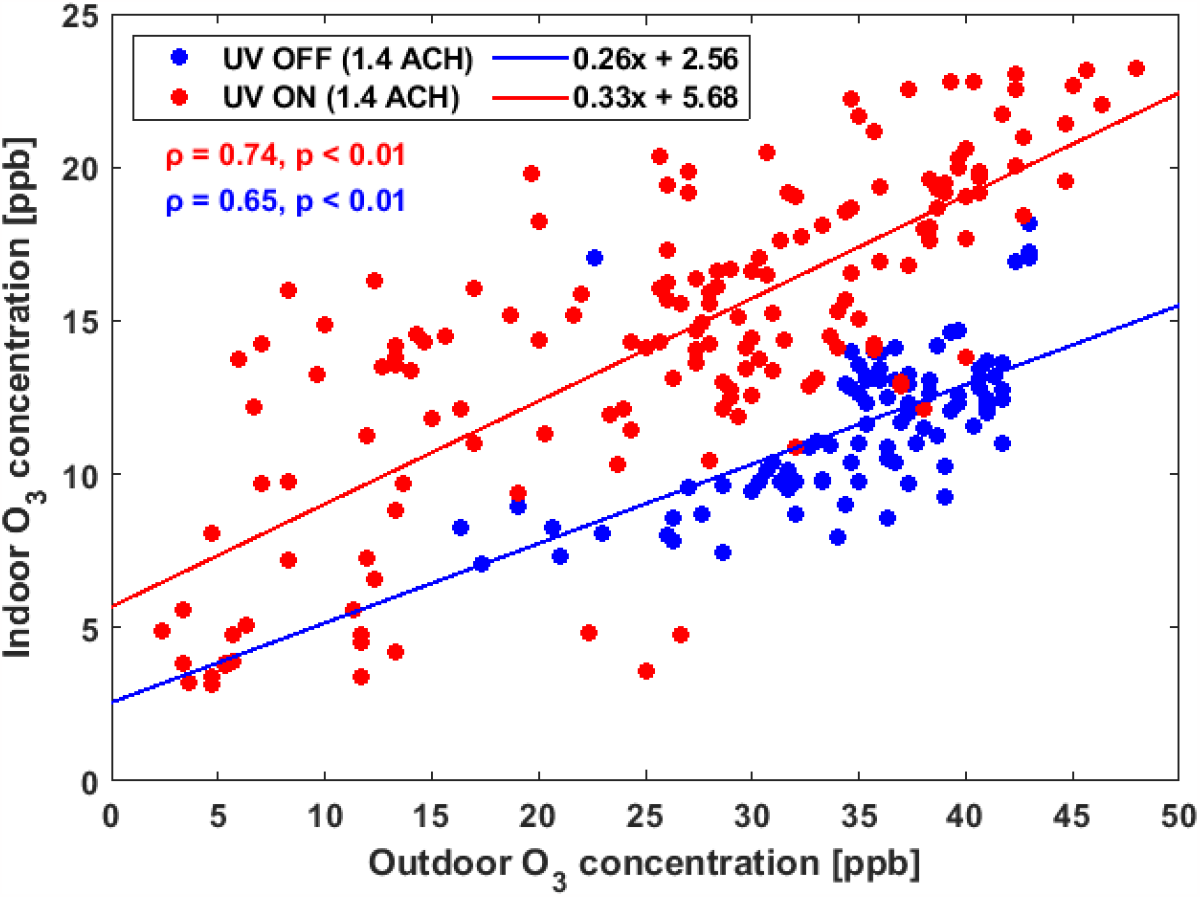
Indoor versus outdoor O_3_ concentrations for the standard hotel room when far-UVC lamps were on and off (with an average air exchange rate of 1.4 ACH). The outdoor O_3_ levels are an average of three Maryland sites (Essex, Glen Burnie, Padonia) in the EPA Ambient Air Monitoring Network. *ρ* is the Spearman’s rank correlation coefficient.

### 3.3. UFP Concentrations

The total UFP concentration in the sealed room ranged from 87 #/cm^3^ to 6262 #/cm^3^ (UV off) and from 110 #/cm^3^ to 3884 #/cm^3^ (UV on) with peaks seemingly unrelated to operation of the far-UVC lamps throughout the sampling period (see Figure 5). The total UFP concentration in the standard room ranged from 167 #/cm^3^ to 2196 #/cm^3^ (UV off) and from 155 #/cm^3^ to 6012 #/cm^3^ (UV on) with major fluctuations also seemingly unrelated to far-UVC usage throughout the sampling period.

UFP concentrations in the sealed room were, on average, lower and less variable than those in the standard room. The concentrations in both rooms were higher during far-UVC usage compared to times when the lights were off (Figure 7). Addition of filtration reduced the particle concentrations in both rooms. Simultaneous measurements of UFP in paired rooms and outdoors were not available.

**Figure 7.**
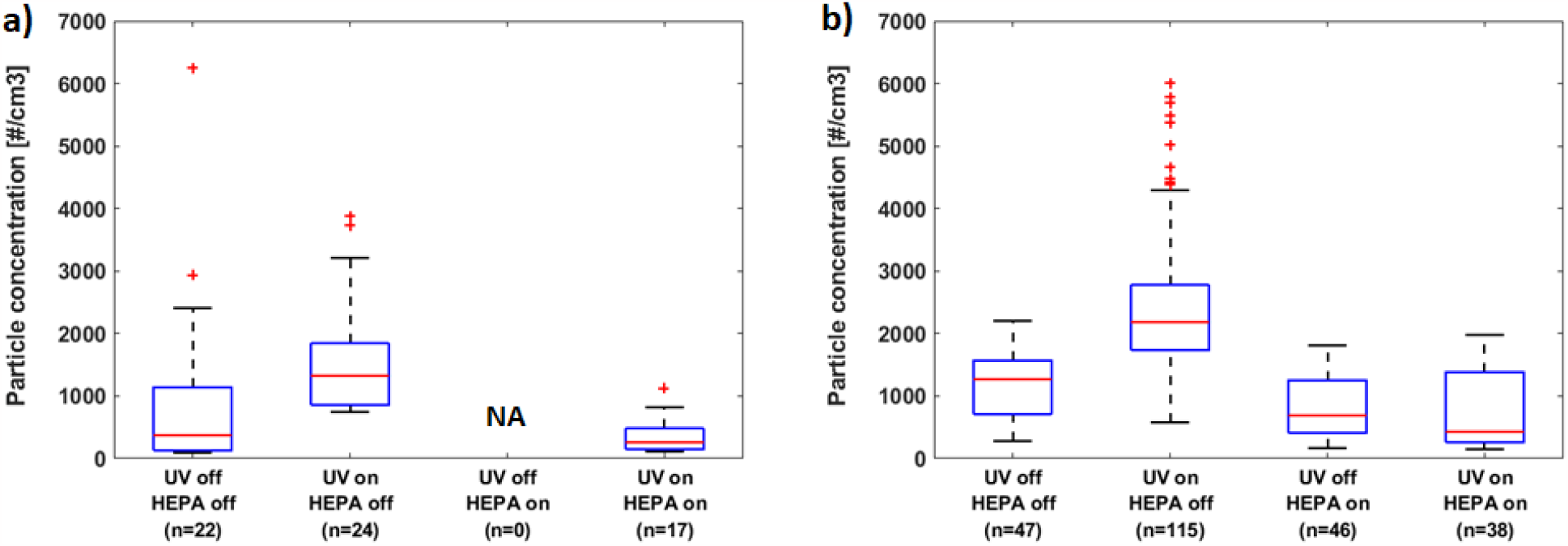
Ultra-fine particle concentrations (17.5 to 289 nm) with and without use of 222 nm far-UVC lamps and HEPA filters a) in the sealed unfurnished room (no data from HEPA on UV off) and b) in the standard room. Boxplots show medians (redline inside the boxes), 25th and 75th percentiles (top and bottom of the boxes), most extreme data not considered outliers (whiskers) and outliers (red crosses) of 1-hr data. n represents the number of 1-hr data points included in each boxplot. HEPA filter units delivered 232 L/s (9.1 ACH) in the sealed room and 116 L/s (7.1 ACH) in the standard room. Simultaneous background measurements to account for indoor and outdoor sources were not collected.

In the standard room, with the far-UVC lamps off, the filtration reduced the average UFP counts from 1173 (95% CI 1023 – 1323) to 807 (95% CI 672 – 943). With the far-UVC lamps on, the filtration reduced the average UFP counts from 2425 (95% CI 2219 – 2630) to 760 (95% CI 563 – 957).

In a simple linear model, without adjustment for indoor or outdoor O_3_, UV in the standard room was associated with an increase in UFP (1019, 95% CI 747 – 1290) and filtration was associated with a decrease in UFP (-1276, 95% CI -1014 to -1537). Further analysis, however, indicated that the relationship between total particle number concentrations, UV, and O_3_ was complex. In the absence of UV, when adjusted for outdoor O_3_ and filtration, increasing indoor O_3_ was associated with increasing UFP concentrations (153 ppb^-1^, p = 3×10^−9^). However, when far-UVC lamps were on and we adjusted for outdoor O_3_ and filtration, indoor O_3_ was associated with decreasing UFP concentrations (-158 ppb^-1^ p = 2×10^−13^). Thus, other factors related to ventilation seemed to have major impacts on UFP concentrations.

## 4. Discussion

### 4.1. O_3_ concentrations

Our findings suggest that 222 nm far-UVC usage increases indoor O_3_ concentrations above levels typically found in buildings. For example, in the absence of indoor sources, indoor O_3_ concentrations (in schools, offices, and residences) are typically around 4-6 ppb^31^. In our study, average O_3_ concentrations in the standard hotel room were 12 ppb with UV off and 14 ppb with UV lamps on. When adjusted for outdoor O_3_, the increase of O_3_ in the standard room during UV usage was estimated to be 5.7 ppb. This increase is similar to the ∼6.5-ppb increase recently reported by Peng et al. (2023)^26^, who measured O_3_ in a sealed office room with and without 222 nm far-UVC lamp usage. All measured indoor O_3_ concentrations in our study and in Peng et al. (2023)^26^ were well below the US ambient (EPA)^32^ and occupational (OSHA)^33^ 8-hour exposure limits (50-100 ppb), and WHO^34^ 8-hour ambient air quality guideline levels (50 ppb). Long-term exposure to ambient O_3_ levels of 30-40 ppb or higher is associated with elevated health risks^35,36^. These levels far exceed the average indoor O_3_ concentrations (with far-UVC lamps on) measured in our study. However, the reader should note that there seems to be no clear threshold for adverse health effects^37^.

The sealed empty hotel room served as a baseline test room in our study, intended to examine worst-case conditions with low air exchange, similar to the experimental conditions in recent publications^23, 26, 27^. Our O_3_ concentration levels in the sealed room were similar to Peng et al. (2023)^26^.

The modelled average fluence rates in this study were 1.7-1.8 μW/cm^2^, which correspond to a 50-51 mJ/cm^2^ dose during an 8-hr period (151-153 mJ/cm^2^ during a 24-h period). These are below ACGIH recommended 8-hr exposure limits (161 mJ/cm^2^ for eyes and 479 mJ/cm^2^ for skin)^38^ but higher than the ICNIRP 8-hr exposure limit^39^ of 23 mJ/cm^2^.

### 4.2. Ultra-fine particle concentrations

In our study, we saw elevated UFP concentrations during far-UVC lamp usage in both rooms. However, linear regression models adjusted for outdoor O_3_ levels indicated that when far-UVC lamps were on, increasing indoor O_3_ was associated with decreasing UFP concentrations in the standard hotel room. This suggests that increases of UFP concentrations in the standard room were mainly influenced by factors other than far-UVC generated O_3_. Local combustion is one potential source of UFPs^40^ that can penetrate indoors (infiltration of outdoor particles). Although doors and windows were closed during our measurements, indoor O_3_ concentrations responded rapidly to outdoor O_3_ changes, suggesting that significant infiltration was occurring. Occasionally noticeable cooking odors, possibly originating from the floor below, suggested presence of significant unmeasured indoor UFP sources. Nevertheless, in-room HEPA filtration produced consistently low UFP concentrations with and without operation of far-UVC lamps.

### 4.3. Limitations of the study

As this study was carried out in a real-world setting, it has limitations that should be considered when assessing the results. One limitation is that we did not carry out local O_3_ and UFP measurements directly outside of the hotel and in the hallways outside of the rooms. Hence, we could not define to what extent the O_3_ or UFP concentration changes were driven by ambient conditions as opposed to far-UVC usage. Also, because the measurements of the different cases (sealed/standard room, UV on/off, etc.) were carried out in series, the data could not be paired. Also, because ambient conditions varied, comparing different cases was not ideal.

We chose to calculate average concentrations (O_3_, UFPs) based on all data in different cases (not only during steady-state conditions) because measurements in real-world settings do not often have steady-state conditions, as was clearly seen in the standard room measurements. This should be kept in mind when interpreting our results, as they might differ from laboratory measurements where it is easier to reach steady-state conditions and use them in the measurements and analysis.

Future studies in real-world settings could benefit from having a control room to monitor different factors simultaneously and form paired data for more accurate assessment of the effect of different factors on O_3_ and UFP formation. It would also be beneficial to monitor O_3_ and UFPs directly outside of the space, since fluctuations in local concentrations might affect the measured values substantially through infiltration.

## 5. Conclusions

222 nm far-UVC is a promising tool for mitigating aerosol spread of infections in high-risk settings, e.g., hotel quarantine facilities. However, recent studies have raised health concerns related to O_3_ production and secondary chemistry caused by the lamps. We found that 222 nm far-UVC lamps do increase O_3_ concentrations indoors, but the average levels are lower than and notably influenced by outdoor concentrations and remain well below EPA 8-hour ambient exposure limits. Additionally, we did not find a clear relationship between UV usage and UFP concentrations. Despite study limitations, our results do not suggest a major impact of far-UVC on indoor UFP concentrations in the real-world environment that we examined. Nevertheless, we should be cognizant of and work to reduce the potential effects far-UVC has on IAQ, especially in environments with low ventilation.

## Data Availability

All data produced in the present study are available upon reasonable request to the authors

## Acknowledgements

The authors would like to thank the Balvi Foundation for funding this study. Also, support from NOAA/ARL, NIST, and MDE for trace gas measurements is gratefully acknowledged.

