## Supporting Information for "Ozone and ultra-fine particle concentrations in a hotel quarantine facility during 222 nm far-UVC air disinfection"

### Modelling of the average fluence rates

The average UV fluence rates in the sealed and standard hotel rooms were modeled with a lighting software (Visual Lighting R2022, Acuity Brands Inc., Atlanta, GA, USA). The model consisted of a simplified room geometry and far-UVC lamps (luminaires), see Figure S1 for illustration of the model and luminaire locations. We had sent similar far-UVC lamps to a third party for goniometry and irradiance measurements. As part of these measurements, we got IES-files that we used in the UV light modelling. The surface reflectance of the UV light was estimated based on values found in the literature^1,2^.

We modelled the fluence rate for both rooms in two different cases: an average fluence rate in the whole room volume and an average value on a horizontal plane 1.75 m above the floor. Both are important for exposure assessment. The volume average estimates the total exposure in the room and the horizontal plane the eye exposure.

The average fluence rate over the total volume of the room was modelled to be 1.8 μW/cm^2^ (8-h dose: 51 mJ/cm^2^) in the sealed room and 1.7 μW/cm^2^ (8-h dose: 50 mJ/cm^2^) standard room. The average fluence rates in the rooms were below ACGIH 8-h limits for eye and skin exposure. In the upper part of the room the average levels were higher than in the total volume: in the horizontal plane 1.75 m above the floor, the average fluence rate was modelled to be 2.8 μW/cm^2^ (80 mJ/cm^2^) in the sealed room and 2.7 μW/cm^2^ (77 mJ/cm^2^) in the standard room. These values do not exceed the ACGIH 8-h limit for eye (161 mJ/cm^2^)^3^ or skin (479 mJ/cm^2^)^3^ but do exceed ICNIRP 8-hr exposure limit^4^ of 23 mJ/cm^2^. For 24-h exposure period the doses were higher than the ACGIH 8-h exposure limit for eyes. It should be noted that the modelled values on the 1.75 m plane might be overestimations since people are not expected to be standing for 8 hours or more, and because the calculation collects flux from all directions whereas e.g., eyes have only limited field of view.

### Ventilation rate measurements and calculations

We monitored ventilation by releasing carbon dioxide (CO_2_) into the rooms at a constant 0.4 L/min rate (GFC171S mass flow controller, Aalborg Instruments & Controls Inc., NY, USA). We used wireless sensors (Aranet4, SAF Tehnika JSC, Latvia) to measure CO_2_ concentrations in the rooms, hallways and outdoors (1-min resolution). The sensors were calibrated with a standard CO_2_ gas with a range of concentrations between 500-10,000 ppm. The sensors also measured ambient temperature, RH, and barometric pressure.

A transient CO_2_ mass balance method^5^ based on a numerical solution was employed to calculate the ventilation rates in the sealed room. A steady-state CO_2_ method^6^ was used to calculate the ventilation rate of the standard room.

**Transient CO_2_ mass balance method:** This method employs a numerical solution to find the CO_2_ concentration for the next time step through the following equation^5^:

$C_{t+1}=6\times{10}^{4} \frac{G}{Q} \left( 1-e^{-\frac{Q}{V}\Delta t} \right)+\left( C_{t}- C_{R} \right)e^{-\frac{Q}{V}\Delta t}+ C_{R}$	(2)

where *C_t_* and *C_R_* are the observed indoor CO_2_ concentration at time *t* and replacement air CO_2_ concentration (ppm), respectively, *G* is the CO_2_ emission rate (L/min), *Q* is the ventilation rate (m^3^/h), *V* is the space volume (m^3^), and *Δt* is the time interval for CO_2_ observations (h). A numerical approach based on the least square method was used to find the ventilation flow rate. The total air exchange rate can be calculated by dividing the ventilation flow rate by the volume of the room, *V* (m^3^). We used this method to calculate the total air exchange rate of the sealed room during the build-up (fan-coil off) and decay phases (fan-coil on). The air exchange rate was found to be 0.1 ACH when the fan-coil unit was on and 0.5 ACH when off.

**Steady-state CO_2_ method:** After indoor CO_2_ concentration reached a steady-state condition, the total ventilation flow rate, *Q* (m^3^/h), was calculated as follows^6^:

$Q=6\times{10}^{4}\frac{G}{C_{S}-C_{R}},$	(1)

where *C_S_* and *C_R_* are indoor and replacement air CO_2_ concentrations (ppm), respectively, *G* is the CO_2_ emission rate from the indoor source (L/min). The total air exchange rate can be calculated by dividing the ventilation flow rate by the volume of the room, *V* (m^3^).

20 steady-state periods were selected to calculate the total air exchange rate in the standard room (see Figure S2). The average air exchange rate was found to be 1.4 ACH.

### Supporting Information figures

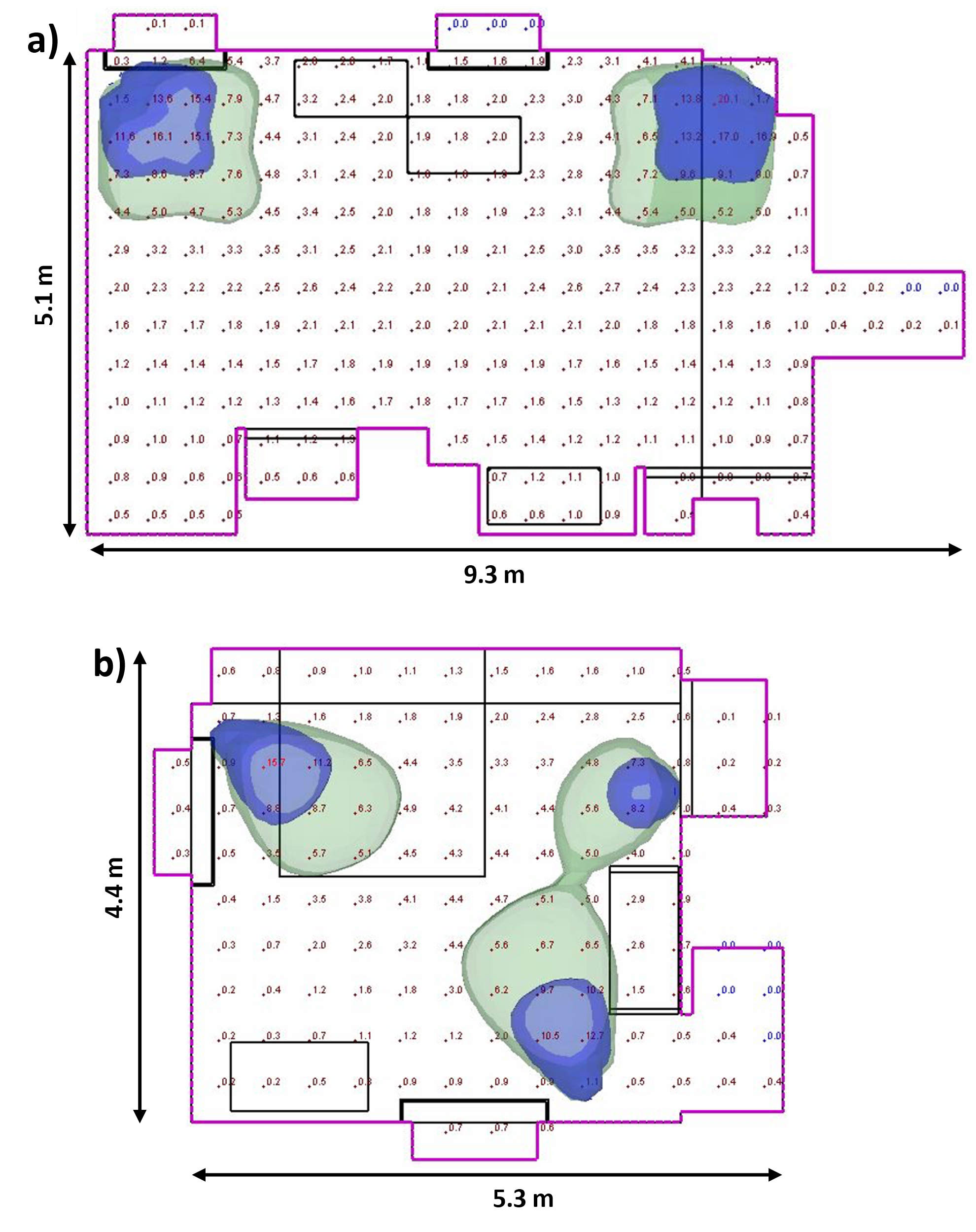

Figure S1. Layout of the simplified room models used in the average fluence rate calculations. a) Sealed room model with the four Ushio B1 modules. b) Standard room model with two Ushio B1 and one B1.5 modules. Blue isosurfaces show where the fluence rate is ≥ 10 μW/cm^2^ and green where the fluence rate is ≥ 5 μW/cm^2^.

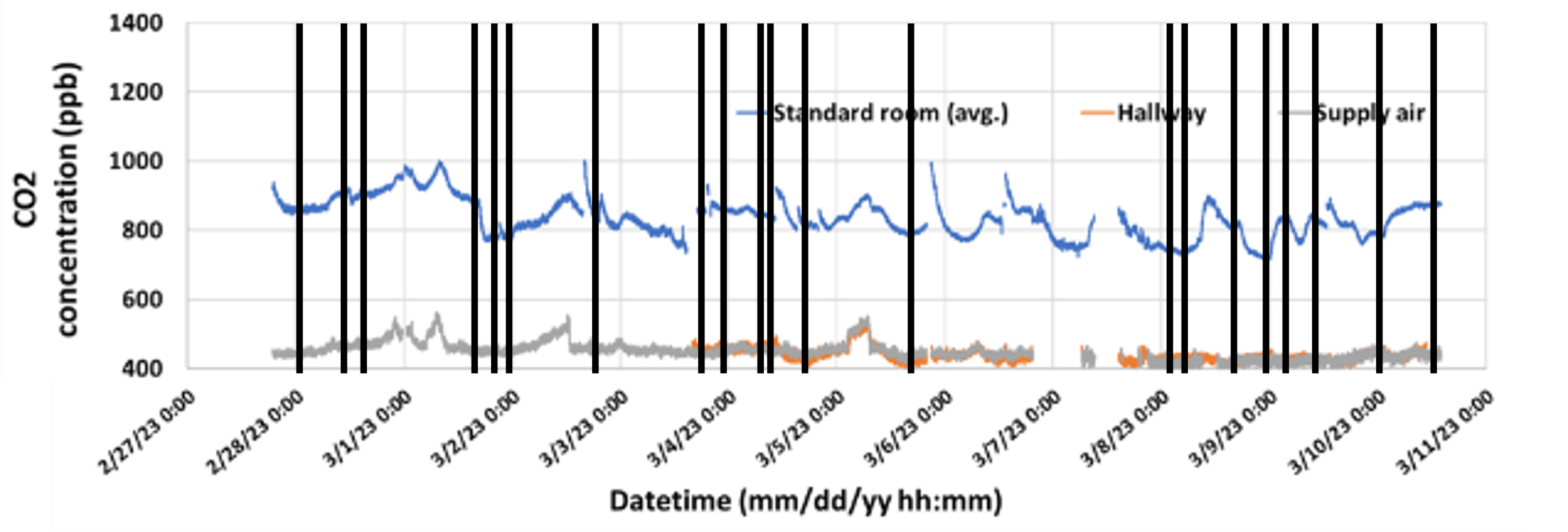

Figure S2. Variation of the CO_2_ concentration (ppm) in the standard room, hallway, and outdoor environment during the experiments. The vertical lines show the time periods selected to calculate the total air exchange rate through the steady-state CO_2_ method.
